# Efficacy and Safety of 5-Day Oral Ensitrelvir for Patients With Mild-to-Moderate COVID-19: The SCORPIO-SR Randomized Clinical Trial

**DOI:** 10.1101/2023.07.11.23292264

**Authors:** Hiroshi Yotsuyanagi, Norio Ohmagari, Yohei Doi, Masaya Yamato, Nguyen Hoang Bac, Bong Ki Cha, Takumi Imamura, Takuhiro Sonoyama, Genki Ichihashi, Takao Sanaki, Yuko Tsuge, Takeki Uehara, Hiroshi Mukae

**Affiliations:** The Institute of Medical Science, The University of Tokyo, Tokyo, Japan; Disease Control and Prevention Center, National Center for Global Health and Medicine, Tokyo, Japan; Division of Infectious Diseases, University of Pittsburgh School of Medicine, Pittsburgh, Pennsylvania, USA; Departments of Microbiology and Infectious Diseases, Fujita Health University School of Medicine, Toyoake, Japan; Infectious Diseases Center, Rinku General Medical Center, Izumisano, Japan; Endoscopic Surgery Training Center, University Medical Center, University of Medicine and Pharmacy, Ho Chi Minh City, Vietnam; Department of Internal Medicine, Chung-Ang Medical Health Care System Hyundae Hospital, Gyeonggi-do, Republic of Korea; Drug Development and Regulatory Science Division, Shionogi & Co., Ltd., Osaka, Japan; Research Division, Shionogi & Co., Ltd., Toyonaka, Japan; Department of Respiratory Medicine, Nagasaki University Graduate School of Biomedical Sciences, Nagasaki, Japan

**Author notes:** CORRESPONDING AUTHOR: Takeki Uehara, Drug Development and Regulatory Science Division, Shionogi & Co., Ltd. Address: 8F, Nissay Yodoyabashi East, 3-3-13 Imabashi, Chuo-ku, Osaka 541-0042, Japan.

## Abstract

**IMPORTANCE:** Treatment options for coronavirus disease 2019 (COVID-19) that can be used irrespective of risk factors for severe disease are warranted.

**OBJECTIVE:** To assess the efficacy and safety of ensitrelvir in patients with mild-to-moderate COVID-19.

**DESIGN:** The phase 3 part of a phase 2/3, double-blind, randomized, placebo-controlled study conducted from February 10 to July 10, 2022.

**SETTING:** A multicenter study conducted at 92 institutions in Japan, Vietnam, and South Korea.

**PARTICIPANTS:** Patients (aged 12 to <70 years) with mild-to-moderate COVID-19 within 120 hours of positive viral testing.

**INTERVENTIONS:** Patients were randomized (1:1:1) to receive once-daily ensitrelvir 125 mg (375 mg on day 1), 250 mg (750 mg on day 1), or placebo for 5 days. Among 1821 randomized patients, 1030 (347, 340, and 343 in the ensitrelvir 125-mg, ensitrelvir 250-mg, and placebo groups, respectively) were randomized in less than 72 hours of disease onset and assessed as the primary analysis population.

**MAIN OUTCOMES AND MEASURES:** The primary end point was the time to resolution of five COVID-19 symptoms (stuffy or runny nose, sore throat, cough, feeling hot or feverish, and low energy or tiredness). Other end points included virologic efficacy and safety.

**RESULTS:** The mean age was 35.7, 35.3, and 34.7 years, and 193 (55.6%), 185 (54.4%), and 174 (50.7%) patients were men in the ensitrelvir 125-mg, ensitrelvir 250-mg, and placebo groups, respectively (intention-to-treat, primary analysis population). A significant difference (P=.04 with a Peto-Prentice generalized Wilcoxon test stratified by vaccination history) was observed in the primary end point between ensitrelvir 125 mg and placebo in the primary analysis population (difference in median, −24.3 hours; 95% confidence interval, −78.7 to 11.7). Viral RNA levels on day 4 and time to negative viral titer demonstrated significant reduction vs placebo. The incidence of adverse events was 44.2%, 53.6%, and 24.8% in the ensitrelvir 125-mg, ensitrelvir 250-mg, and placebo groups, respectively. No treatment-related serious adverse events were reported.

**CONCLUSIONS AND RELEVANCE:** Treatment with ensitrelvir 125 mg demonstrated clinical and antiviral efficacy without new safety concerns. Generalizability to non-Asian populations should be confirmed.

**TRIAL REGISTRATION:** Japan Registry of Clinical Trials identifier: <u>jRCT2031210350</u>.

**Key Points:** *Question:* Can ensitrelvir, an oral severe acute respiratory syndrome coronavirus 2 3C-like protease inhibitor, shorten the duration of symptoms in patients with mild-to-moderate COVID-19 irrespective of risk factors for severe disease?

*Findings:* In this phase 3 part of a phase 2/3, double-blind, randomized study SCORPIO-SR, a statistically significant difference was observed in the time to resolution of five COVID-19 symptoms between ensitrelvir 125 mg and placebo in patients randomized in less than 72 hours of disease onset. Viral RNA and viral titer demonstrated significant reduction vs placebo.

*Meaning:* Ensitrelvir 125 mg treatment shortened time to resolution of key COVID-19 symptoms.

## Introduction

Coronavirus disease 2019 (COVID-19) has rapidly spread worldwide, and in November 2021, the severe acute respiratory syndrome coronavirus 2 (SARS-CoV-2) omicron variant was declared a variant of concern by the World Health Organization.^1^ Patients infected with the omicron variant generally experience mild symptoms.^2–4^ However, despite the worldwide administration of vaccinations, the omicron BA.5* subvariant, which evades vaccine-induced immunity, has become dominant and has resulted in a rapid increase in new COVID-19 cases.^5^ Moreover, emergence of the omicron XBB* and BQ.1* sublineages, which also evade immunity, has been reported worldwide.^6^

Several antiviral drugs against SARS-CoV-2 infection are available worldwide, such as RNA polymerase inhibitors remdesivir^7^ and molnupiravir,^8^ and a SARS-CoV-2 3C-like (3CL) protease inhibitor nirmatrelvir combined with pharmacokinetic booster ritonavir.^9^ However, their clinical trials were conducted in high-risk, unvaccinated patients and at a time when the SARS-CoV-2 Delta or pre-Delta variants were predominant. Therapeutic monoclonal antibodies against SARS-CoV-2 have also been used for COVID-19, but recent *in vitro* studies suggest that most of them are less efficacious to omicron subvariants than to previous variants.^10–12^ Novel treatment options that can be used irrespective of risk factors for severe disease are thus warranted.

Ensitrelvir fumaric acid is an oral SARS-CoV-2 3CL protease inhibitor that originated through collaborative research between Shionogi & Co., Ltd. and Hokkaido University in Japan.^13^ Ensitrelvir treatment showed antiviral effects against multiple SARS-CoV-2 variants of concern, including omicron subvariants, in *in vitro* and *in vivo* studies.^13–17^ A seamless, phase 2/3, double-blind, randomized, placebo-controlled study is currently underway (Japan Registry of Clinical Trials identifier: jRCT2031210350). In the phase 2a and 2b parts, ensitrelvir treatment demonstrated decreased viral load vs placebo.^18,19^ Moreover, improvements in four respiratory symptoms (stuffy or runny nose, sore throat, shortness of breath, and cough) and the composite of the four respiratory symptoms and feverishness were shown in the phase 2b part.^19^ The SCORPIO-SR trial is the phase 3 part of the phase 2/3 study and assessed the efficacy and safety of 5-day, once-daily ensitrelvir in patients with mild-to-moderate COVID-19.

## Methods

### Study Design

The SCORPIO-SR trial was conducted from February 10 to July 10, 2022, across 92 institutions in Japan (65 sites), Vietnam (2 sites), and South Korea (25 sites). The study protocol has been published previously.^20^ Patients with mild-to-moderate COVID-19 were randomized (1:1:1) to the ensitrelvir 125 mg, ensitrelvir 250 mg, or placebo group. The patients were followed up until day 28. Considering that the pharmacokinetic/pharmacodynamic analysis of phase 2b part of the current phase 2/3 study did not show a clear difference in antiviral efficacy between the two ensitrelvir dose groups, the 125-mg group was chosen as the efficacy analysis population. Analyses between the ensitrelvir 250-mg and placebo groups were performed as a secondary comparison.^20^

The current phase 2/3 study employed a seamless design to maximize patient recruitment during the SARS-CoV-2 epidemic, and the current phase 3 part was initiated with continuous patient enrollment after the phase 2b part. The phase 3 protocol was amended based on data obtained in the phase 2b part and in view of the changes in clinical features of the epidemic caused by the omicron variant and its subvariants. All protocol amendments, including the changes in the primary endpoint, analysis populations, statistical tests, and sample size estimation, were made blinded from the phase 3 data and were finalized based on consultations with infectious disease experts and regulatory authorities.^20^

This study was conducted in accordance with the principles of the Declaration of Helsinki, Good Clinical Practice guidelines, and other applicable laws and regulations. The study protocol was reviewed and approved by the institutional review boards of all the participating institutions (see the List of Investigators in the Supplement). Written informed consent was obtained from all patients or their legally acceptable representatives.

### Patients

Eligible patients with mild-to-moderate COVID-19 were those aged 12 to <70 years who tested positive for SARS-CoV-2 within 120 hours prior to randomization. Patients should have had a time of ≤120 hours from the onset of COVID-19 symptoms to randomization and at least one moderate or severe symptom or worsening of an existing moderate or severe symptom among the 12 COVID-19 symptoms defined based on the United States Food and Drug Administration (FDA) guidance^21^ (eMethods). Key exclusion criteria included an awake oxygen saturation of ≤93% (room air), supplemental oxygen requirement, anticipated COVID-19 exacerbation within 48 hours of randomization, suspected active and systemic infections other than COVID-19 requiring treatment, current or chronic history of moderate or severe liver disease, known hepatic or biliary abnormalities (except for Gilbert’s syndrome or asymptomatic gallstones), and moderate-to-severe kidney disease (eMethods).

### Randomization, Blinding, and Treatment

Patient randomization was performed using an interactive response technology system and stratification by the time from the onset of COVID-19 to randomization (<72 hours vs ≥72 hours) and SARS-CoV-2 vaccination history (at least one dose of vaccination: yes vs no). All patients and study staff were blinded to the treatment, and emergency unblinding per the investigator’s request was allowed in case of occurrence of adverse events to determine the appropriate therapy for the patient. Patients received ensitrelvir (375 mg on day 1 and 125 mg on days 2 through 5 or 750 mg on day 1 and 250 mg on days 2 through 5) or matching placebo tablets, which were identical in appearance and packaging, without dose modification. Treatment was discontinued in case of COVID-19 exacerbation, serious or intolerable adverse events, and pregnancy (eMethods).

### End Points

The primary end point was the time to resolution of five COVID-19 symptoms (stuffy or runny nose, sore throat, cough, feeling hot or feverish, and low energy or tiredness) on the basis that they were most commonly observed symptoms in the phase 2b part of the current phase 2/3 study, conducted during the omicron BA.1 epidemic.^20^ Key secondary end points were the change from baseline in SARS-CoV-2 viral RNA level on day 4 (key secondary end point 1) and time to first negative SARS-CoV-2 viral titer (key secondary end point 2). The time to resolution of 12 COVID-19 symptoms (above-mentioned five symptoms, shortness of breath, muscle or body aches, headache, chills or shivering, nausea, vomiting, and diarrhea) and 14 COVID-19 symptoms (above-mentioned 12 symptoms, anosmia, and dysgeusia) and the SARS-CoV-2 viral RNA and viral titer up to day 21 were assessed as other secondary end points. Resolution of five, 12, and 14 COVID-19 symptoms was achieved when all the symptoms disappeared, improved, or maintained after first administration of study intervention and the symptom resolution was sustained for at least 24 hours in a patient. Each of the 12 COVID-19 symptoms was rated on a 4-point scale (None=0, Mild=1, Moderate=2, or Severe=3), whereas anosmia and dysgeusia were rated on a 3-point scale (Same as usual=0, Less than usual=1, or No sense of smell/taste=2) by the patient (eMethods). The safety end point was the incidence of adverse events that emerged after treatment initiation.

### Assessments

The severity of each COVID-19 symptom was recorded in a diary twice daily (morning and evening) until day 9 and once daily (evening) from days 10 to 21. SARS-CoV-2 viral titer and viral RNA were quantified using nasopharyngeal swabs collected on days 1 (before drug administration), 2 to 6 (days 3 and 5 as optional visits), 9, 14, and 21 (or study discontinuation). Adverse events were coded using the Medical Dictionary for Regulatory Activities, version 24.0 (eMethods).

### Statistical Analysis

Considering the main pathophysiology of COVID-19 (viral proliferation for several days followed by inflammatory reaction by the host immune system starting around 7 days after onset), antiviral treatment is expected to be most effective when initiated in the early phase of infection.^20^ Based on an epidemiologic analysis in Japan, 62% of patients with COVID-19 hospitalized during the omicron BA.5 epidemic experienced disease progression within 3 days of the onset.^22^ To evaluate the antiviral and clinical efficacy of ensitrelvir in the optimal patient population, the primary analysis population for the primary and key secondary end points was defined as patients randomized in less than 72 hours of disease onset in the intention-to-treat population.^20^ As reported previously,^20^ assuming a Weibull distribution in the time to resolution of the five COVID-19 symptoms, 230 patients per group were required to compare the survival distributions of the primary end point between the ensitrelvir 125-mg and placebo groups using the Peto-Prentice generalized Wilcoxon test with 80% power and a two-sided significance level of .05. Assuming a 10% dropout rate before enrollment, the final sample size for the primary analysis population was set to 260 patients per group (780 patients in total).^20^ Of note, patient enrollment was continued as originally planned to include 595 patients randomized within 120 hours of disease onset each in the ensitrelvir 125-mg, 250-mg, and placebo groups.

The statistical significance vs placebo for the primary end point in the primary analysis population was tested using a Peto-Prentice generalized Wilcoxon test stratified by SARS-CoV-2 vaccination history (yes or no). A fixed-sequence hierarchical testing procedure was applied to control for type I errors. The test was performed in the order of primary end point, key secondary end point 1, and key secondary end point 2 in the primary analysis population. The tests were repeated in the same order in patients randomized within 120 hours of disease onset until failure in statistical significance (two-sided significance level of ≥.05).^20^ All statistical comparisons were performed at a two-sided significance level of .05. No imputation was performed for missing data. All analyses were performed using SAS version 9.4 (SAS Institute, Inc, Cary, NC, USA; eMethods).

## Results

### Patients

A total of 1821 patients were enrolled; 607, 606, and 608 were randomized to the ensitrelvir 125-mg, ensitrelvir 250-mg, and placebo groups, respectively (Figure 1). Of the 1821 patients, 1769 (97.1%) completed the study and 1030 (56.6%) were randomized in less than 72 hours of disease onset.

**Figure 1.**
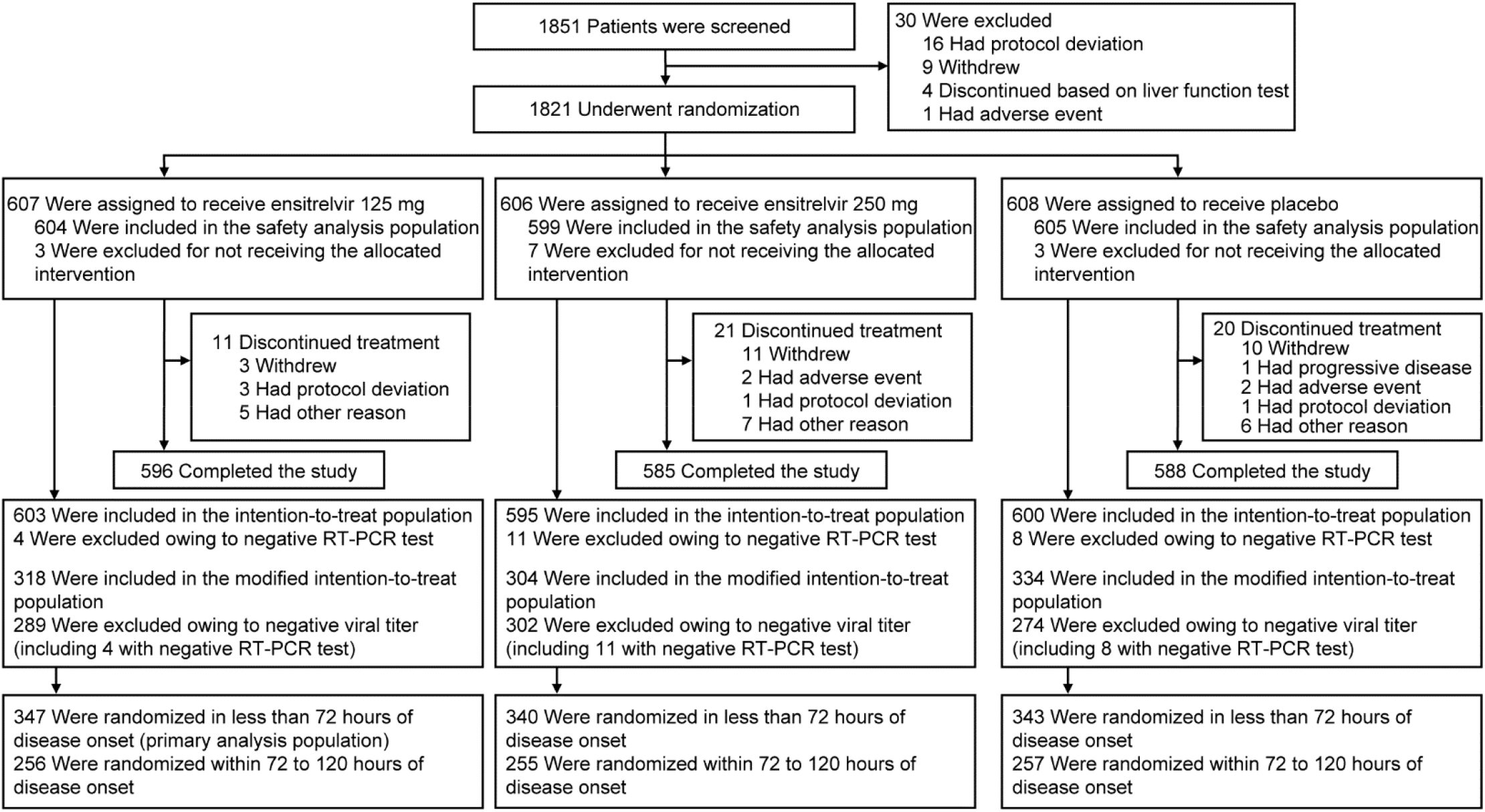
Patient Disposition. The intention-to-treat population comprised all patients who tested positive for SARS-CoV-2 viral RNA at baseline, as confirmed by an RT-PCR test based on the nasopharyngeal swab sample. The modified intention-to-treat population comprised all patients who tested positive for SARS-CoV-2 viral RNA and who had detectable SARS-CoV-2 viral titer at baseline. RT-PCR, reverse transcription-polymerase chain reaction; SARS-CoV-2, severe acute respiratory syndrome coronavirus 2.

Patient characteristics were similar across the groups (Table 1 and eTable 1) and were largely representative of the expected patient population (eTable 2). In all groups, the mean age was approximately 35 years, approximately half of the patients were men, and almost all patients were Asian. The majority (>90%) of patients had received ≥2 doses of the SARS-CoV-2 vaccine. Prior acetaminophen use was recorded for approximately 26% to 36% of the patients. Most (≥85%) patients were infected with the omicron BA.1 or BA.2 subvariant. The most common COVID-19 symptoms across treatment groups were stuffy or runny nose, sore throat, cough, feeling hot or feverish, and low energy or tiredness (Table 1). Approximately 70% of the patients were those without risk factors for severe disease (Table 1 and eTable 3).

**Table 1.**
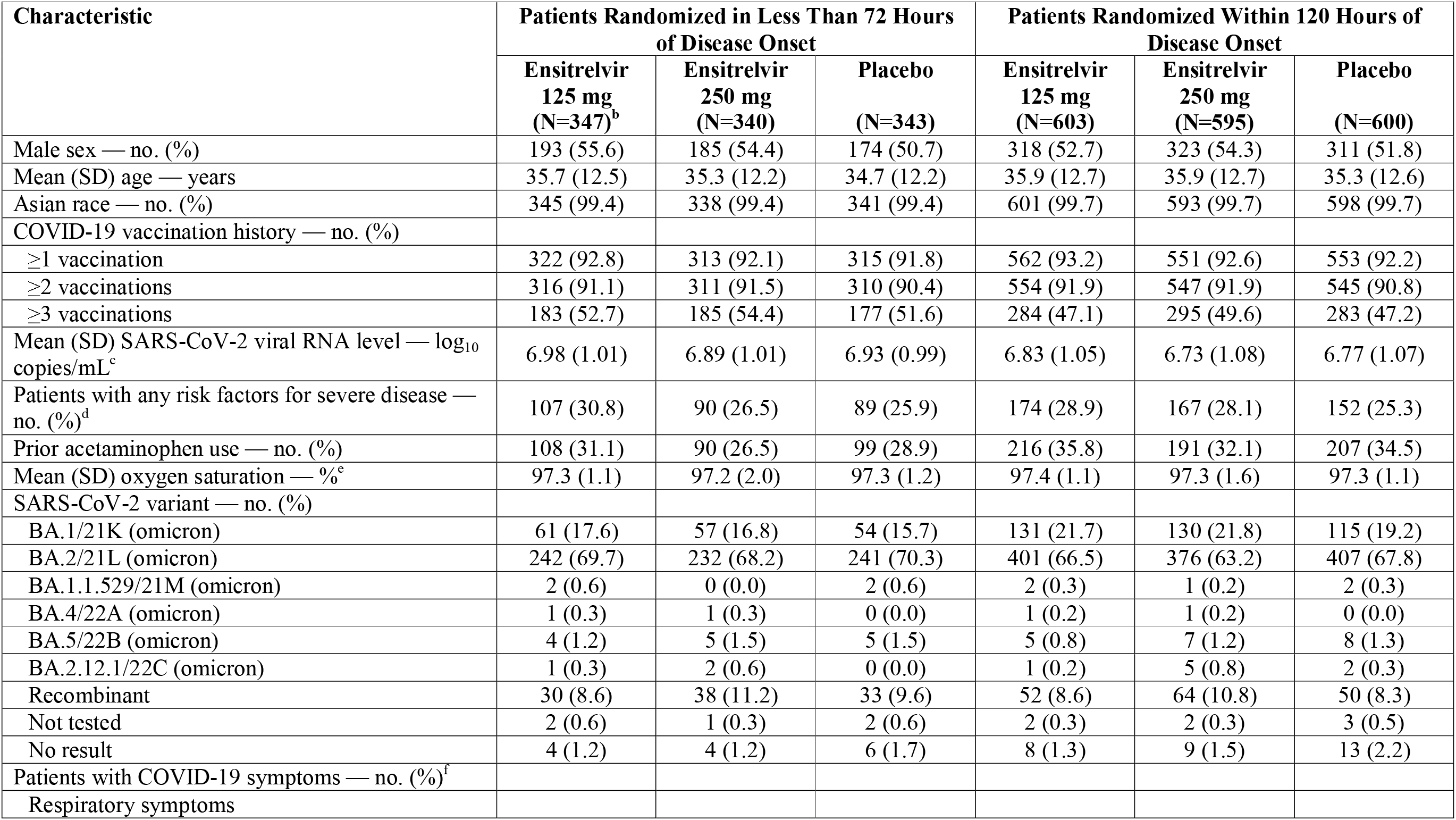
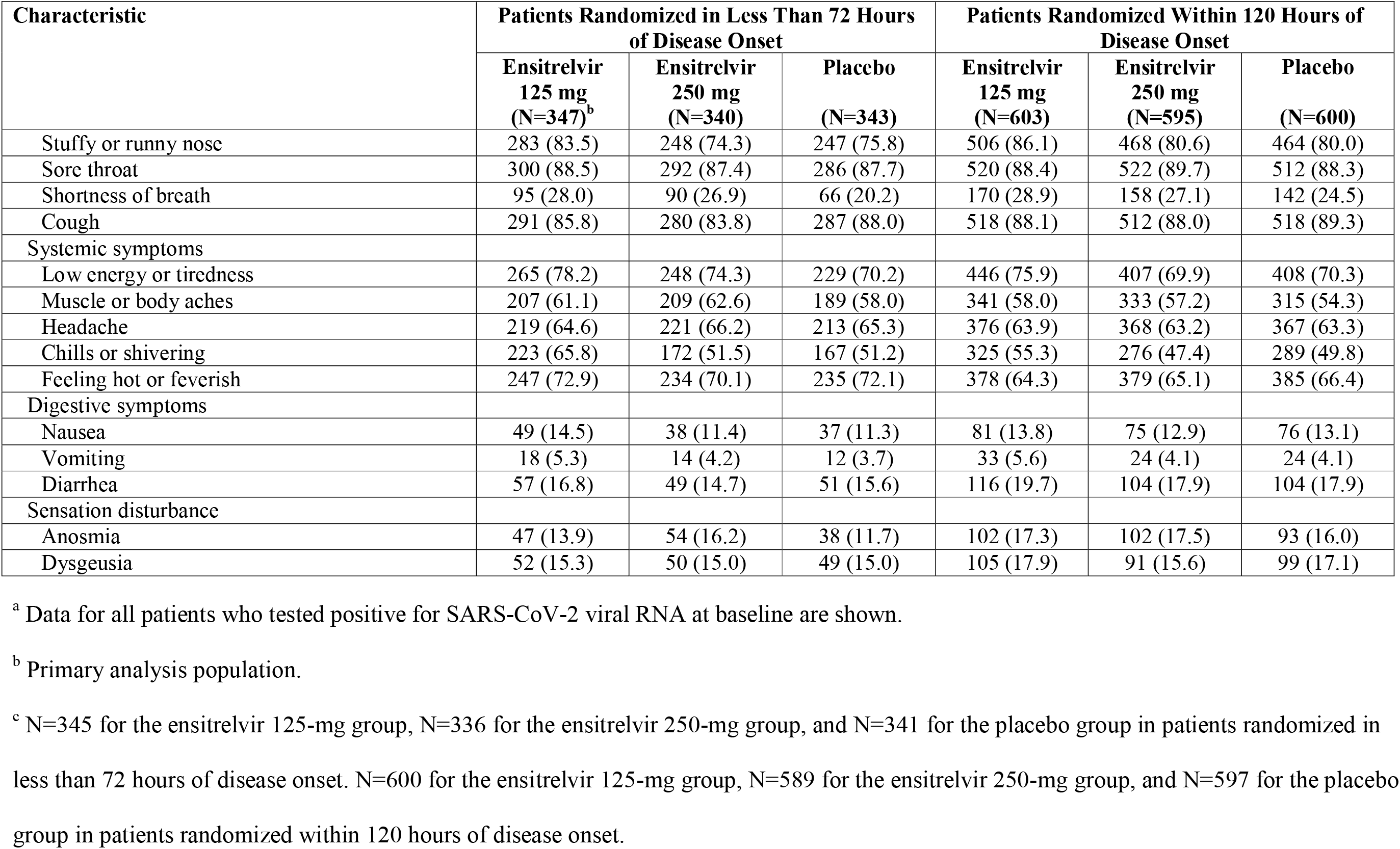
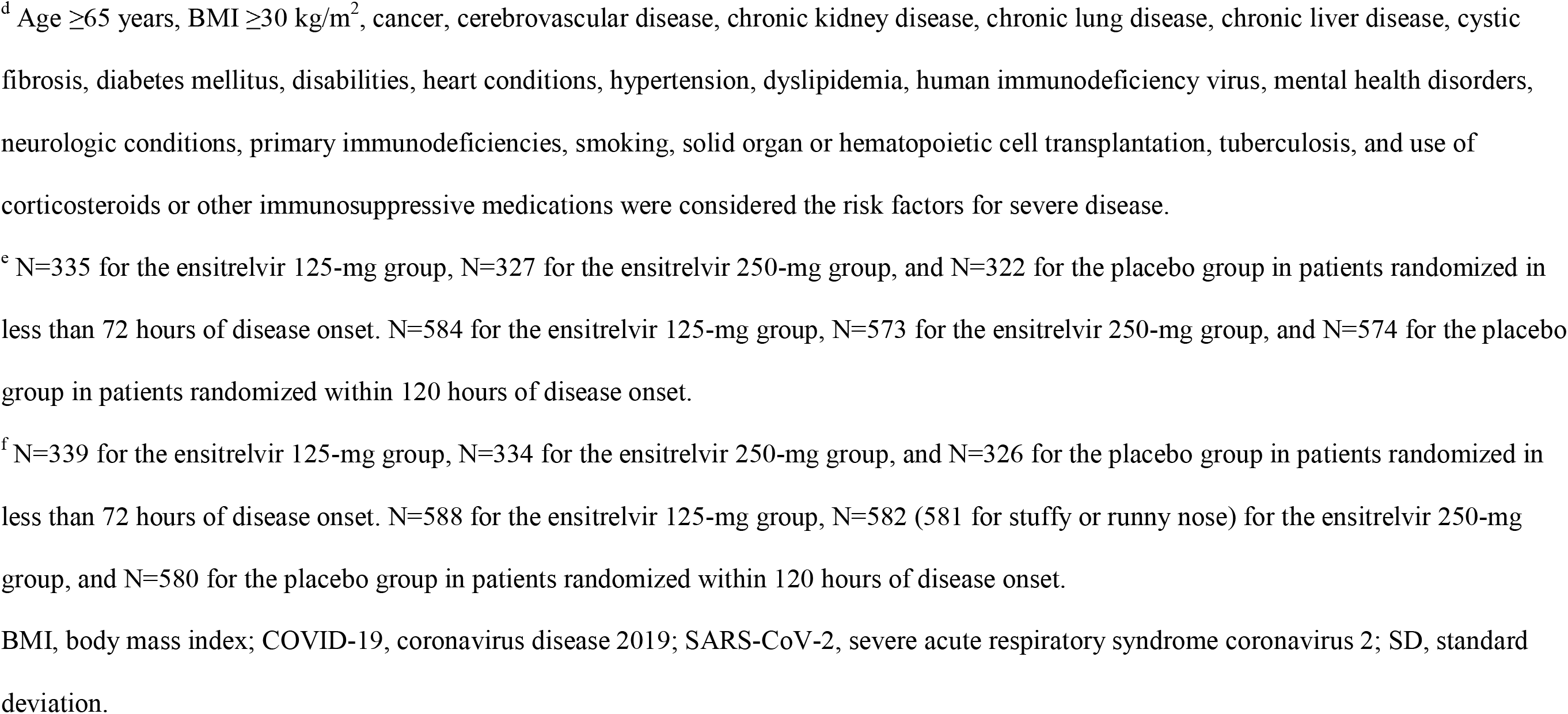
Demographic and Clinical Characteristics of the Patients at Baseline (Intention-to-Treat Population)^a^.

### Primary End Point

A significant difference (P=.04 with a Peto-Prentice generalized Wilcoxon test) was observed between ensitrelvir 125 mg (median, 167.9 hours; 95% confidence interval [CI], 145.0 to 197.6) and placebo (median, 192.2 hours; 95% CI, 174.5 to 238.3) in the time to resolution of the five COVID-19 symptoms among patients randomized in less than 72 hours of disease onset, i.e., primary analysis population (difference in median, −24.3 hours; 95% CI, −78.7 to 11.7; Figure 2A). No significant difference in the time to resolution of the five COVID-19 symptoms was observed between ensitrelvir 125 mg and placebo for patients randomized within 120 hours of disease onset (Figure 2B).

**Figure 2.**
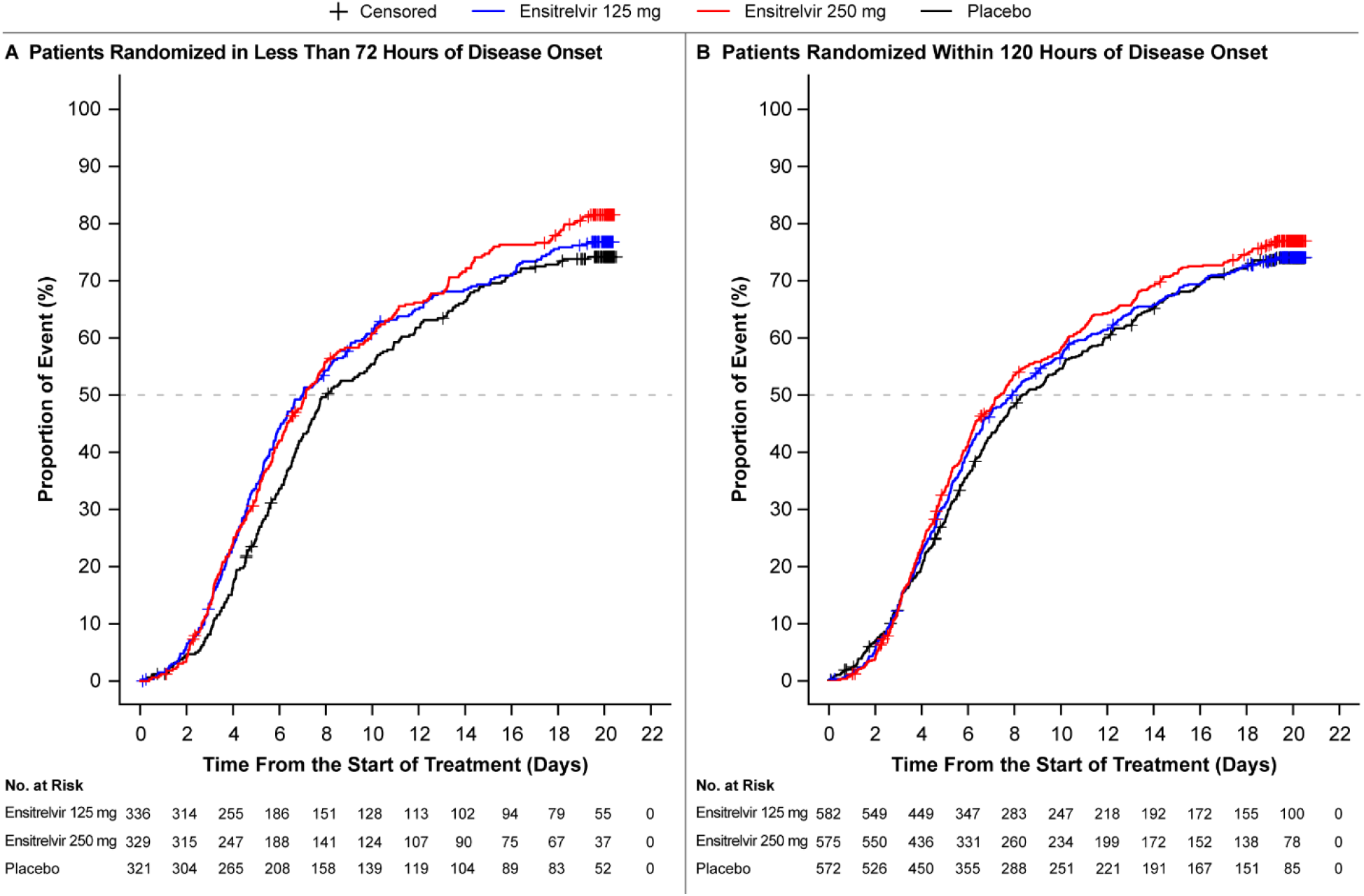
Time to Resolution of Five COVID-19 Symptoms (Intention-to-Treat Population) The analysis was performed for all patients who tested positive for SARS-CoV-2 viral RNA at baseline. Patients randomized in less than 72 hours of disease onset in the ensitrelvir 125-mg group were defined as the primary analysis population. A Peto-Prentice generalized Wilcoxon test was applied to test the statistical significance vs placebo. The test was stratified by SARS-CoV-2 vaccination history (yes or no) for patients randomized in less than 72 hours (panel A) and time from onset to randomization (<72 hours or ≥72 hours) and SARS-CoV-2 vaccination history (yes or no) for patients randomized within 120 hours (panel B). The five COVID-19 symptoms were stuffy or runny nose, sore throat, cough, feeling hot or feverish, and low energy or tiredness. Among patients randomized within 120 hours of disease onset (panel B), the median difference in the time to resolution of the five COVID-19 symptoms between ensitrelvir 125 mg and placebo was −10.6 hours (95% confidence interval, −56.9 to 21.3). COVID-19, coronavirus disease 2019; RNA, ribonucleic acid; SARS-CoV-2, severe acute respiratory syndrome coronavirus 2.

### Key Secondary End Points

The least squares mean change from baseline in SARS-CoV-2 viral RNA level (log_10_ copies/mL) on day 4 in the primary analysis population was −2.48 (standard error [SE], 0.08) with the ensitrelvir 125-mg group, as compared with −1.01 (SE, 0.08) with the placebo group (P<.001) (Table 2). A significant difference (P<.001) was observed in the time to first negative SARS-CoV-2 viral titer in the primary analysis population between ensitrelvir 125 mg (median, 36.2 hours; 95% CI, 23.4 to 43.2) and placebo (median, 65.3 hours; 95% CI, 62.0 to 66.8; eFigure 1A). Similar results were obtained for patients randomized within 120 hours of disease onset (eFigure 1B).

**Table 2.**
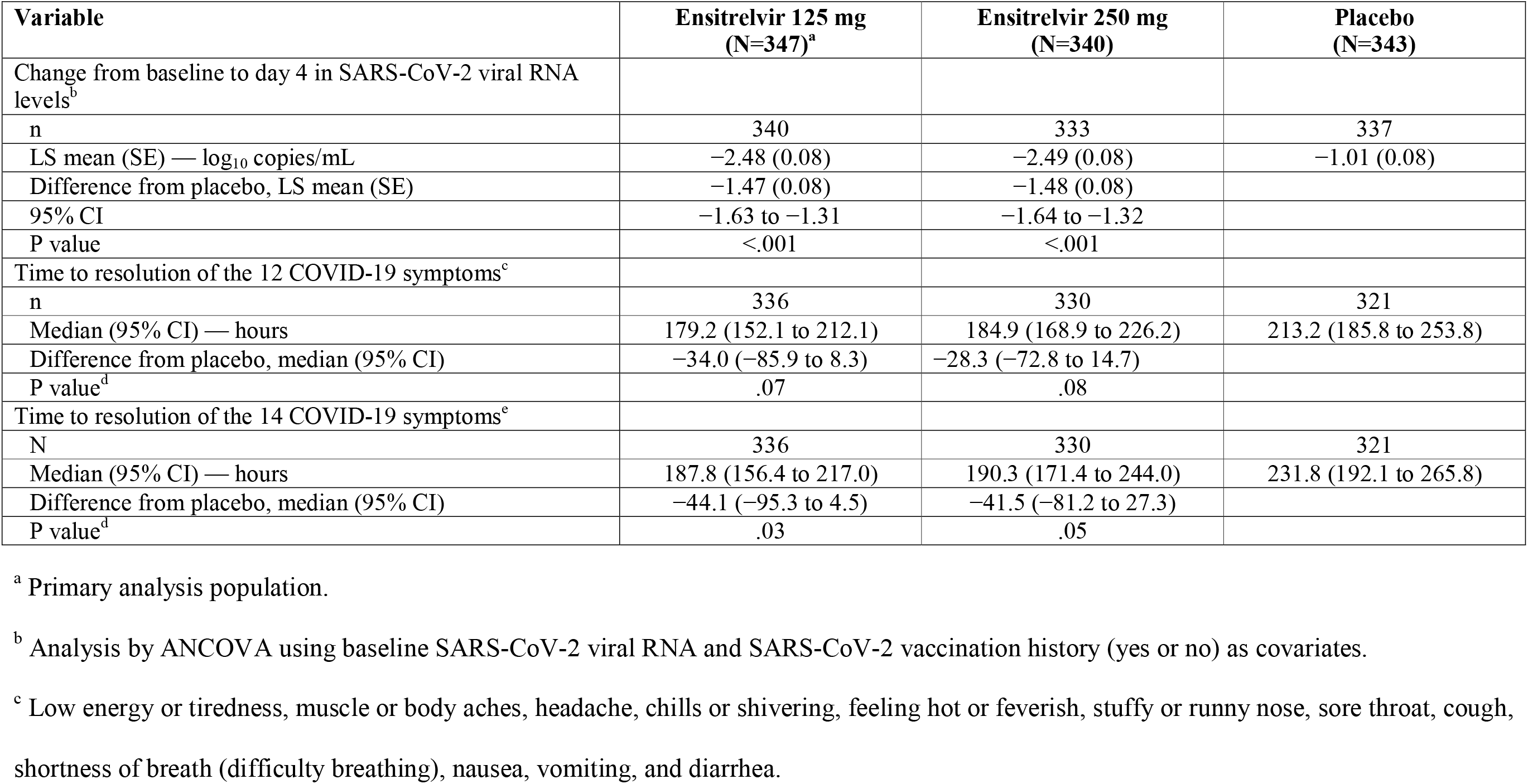
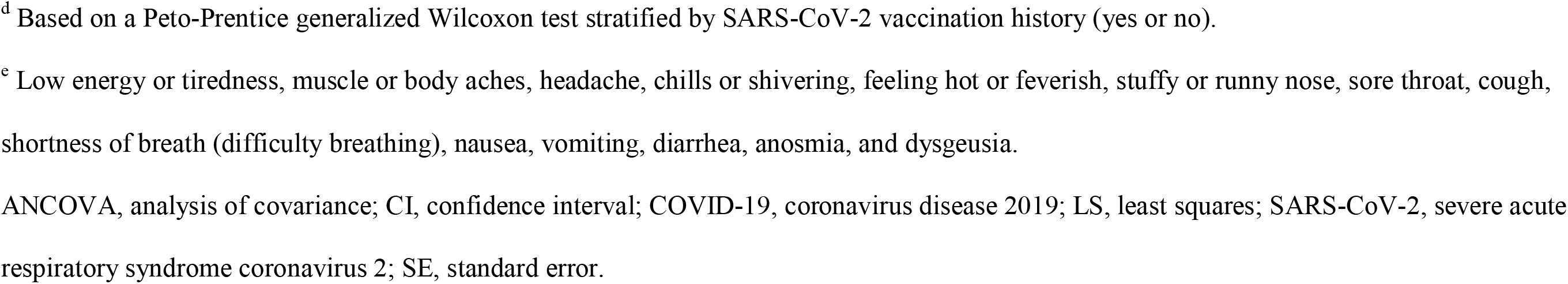
Change from Baseline to Day 4 in SARS-CoV-2 Viral RNA Levels, Time to Resolution of the 12 COVID-19 Symptoms, and Time to Resolution of the 14 COVID-19 Symptoms (Intention-to-Treat Population, Patients Randomized in Less Than 72 Hours of Disease Onset)

| Variable | Ensitrelvir 125 mg<br>(N=347)<sup>a</sup> | Ensitrelvir 250 mg<br>(N=340) | Placebo<br>(N=343) |
| --- | --- | --- | --- |
| Change from baseline to day 4 in SARS-CoV-2 viral RNA levels<sup>b</sup> |  |  |  |
| n | 340 | 333 | 337 |
| LS mean (SE) — log<sub>10</sub> copies/mL | −2.48 (0.08) | −2.49 (0.08) | −1.01 (0.08) |
| Difference from placebo, LS mean (SE) | −1.47 (0.08) | −1.48 (0.08) |  |
| 95% CI | −1.63 to −1.31 | −1.64 to −1.32 |  |
| P value | <.001 | <.001 |  |
| Time to resolution of the 12 COVID-19 symptoms<sup>c</sup> |  |  |  |
| n | 336 | 330 | 321 |
| Median (95% CI) — hours | 179.2 (152.1 to 212.1) | 184.9 (168.9 to 226.2) | 213.2 (185.8 to 253.8) |
| Difference from placebo, median (95% CI) | −34.0 (−85.9 to 8.3) | −28.3 (−72.8 to 14.7) |  |
| P value<sup>d</sup> | .07 | .08 |  |
| Time to resolution of the 14 COVID-19 symptoms<sup>c</sup> |  |  |  |
| N | 336 | 330 | 321 |
| Median (95% CI) — hours | 187.8 (156.4 to 217.0) | 190.3 (171.4 to 244.0) | 231.8 (192.1 to 265.8) |
| Difference from placebo, median (95% CI) | −44.1 (−95.3 to 4.5) | −41.5 (−81.2 to 27.3) |  |
| P value<sup>d</sup> | .03 | .05 |  |

<sup>a</sup> Primary analysis population.
<sup>b</sup> Analysis by ANCOVA using baseline SARS-CoV-2 viral RNA and SARS-CoV-2 vaccination history (yes or no) as covariates.
<sup>c</sup> Low energy or tiredness, muscle or body aches, headache, chills or shivering, feeling hot or feverish, stuffy or runny nose, sore throat, cough, shortness of breath (difficulty breathing), nausea, vomiting, and diarrhea.
<sup>d</sup> Based on a Peto-Prentice generalized Wilcoxon test stratified by SARS-CoV-2 vaccination history (yes or no).
<sup>e</sup> Low energy or tiredness, muscle or body aches, headache, chills or shivering, feeling hot or feverish, stuffy or runny nose, sore throat, cough, shortness of breath (difficulty breathing), nausea, vomiting, diarrhea, anosmia, and dysgeusia.
ANCOVA, analysis of covariance; CI, confidence interval; COVID-19, coronavirus disease 2019; LS, least squares; SARS-CoV-2, severe acute respiratory syndrome coronavirus 2; SE, standard error.

### Other Secondary End Points

No significant difference was observed in the time to resolution of the 12 COVID-19 symptoms in the primary analysis population between ensitrelvir 125 mg and placebo. A significant difference (P=.03) was observed in the time to resolution of the 14 COVID-19 symptoms in the primary analysis population between ensitrelvir 125 mg (median, 187.8 hours; 95% CI, 156.4 to 217.0) and placebo (median, 231.8 hours; 95% CI, 192.1 to 265.8; Table 2).

No significant difference from placebo was observed with ensitrelvir 125 mg in the time to resolution of the 12 and 14 COVID-19 symptoms in patients randomized within 120 hours of disease onset (eTable 4). SARS-CoV-2 viral RNA levels and SARS-CoV-2 viral titers in the ensitrelvir 125-mg group were significantly lower than those in the placebo group up to day 9 and day 6, respectively, irrespective of time to randomization (eFigure 2A-C and eFigure 3A-C).

### Safety

The incidence of adverse events was 44.2%, 53.6%, and 24.8% in the ensitrelvir 125-mg, ensitrelvir 250-mg, and placebo groups, respectively; the incidence of treatment-related adverse events was 24.5%, 36.2%, and 9.9% in the ensitrelvir 125-mg, ensitrelvir 250-mg, and placebo groups, respectively (Table 3). Most of the adverse events were mild, and a majority of treatment-related adverse events resolved without sequelae.

**Table 3.**
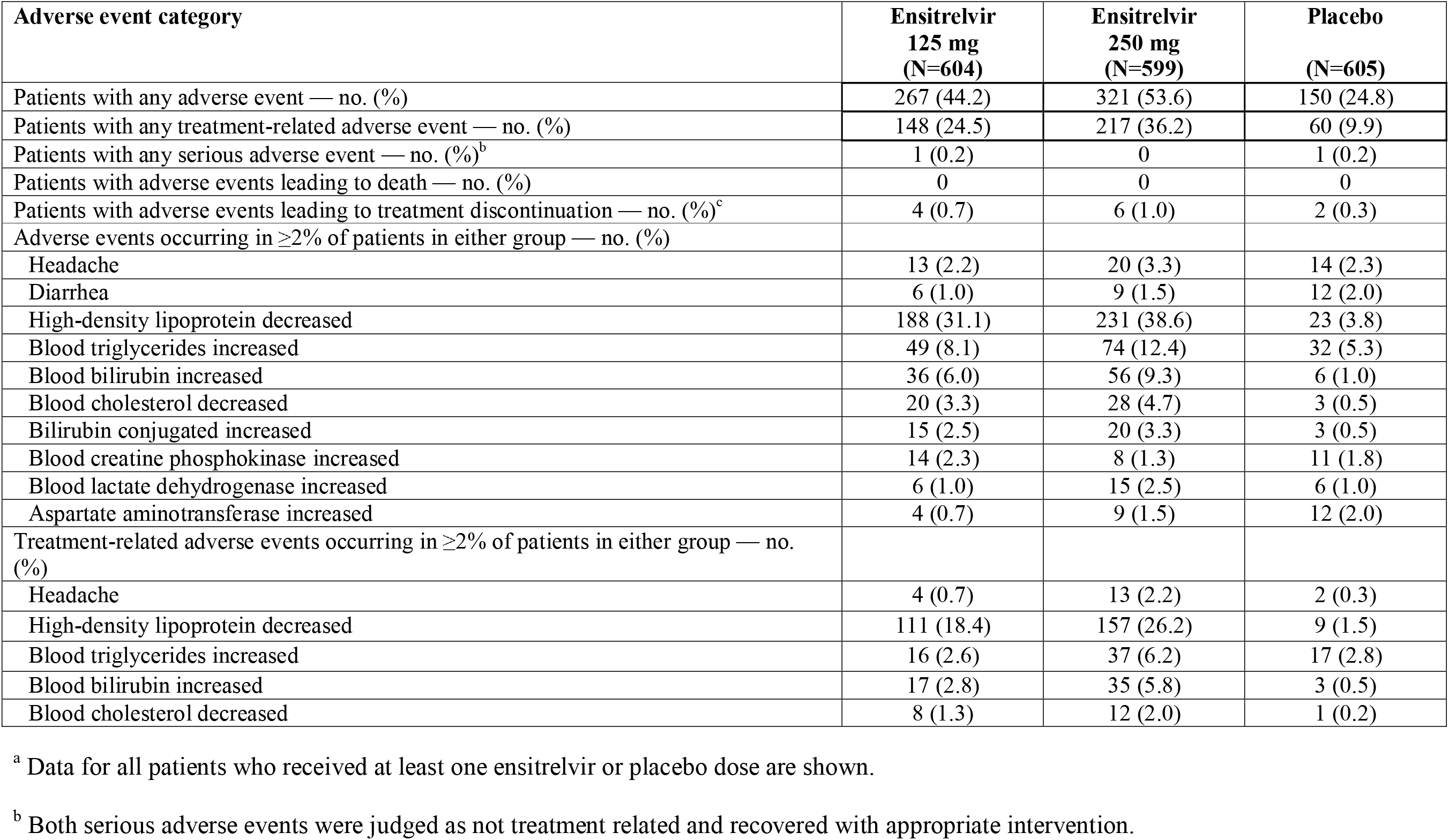
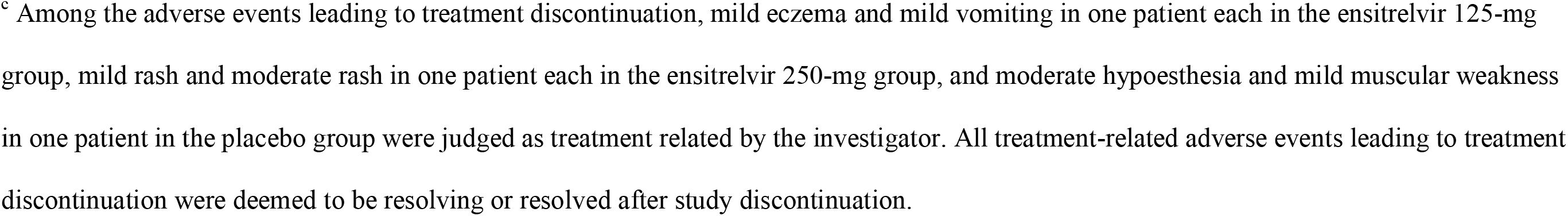
Summary of Adverse Events (Safety Analysis Population)^a^.

| **Adverse event category** | **Ensitrelvir**<br>**125 mg**<br>**(N=604)** | **Ensitrelvir**<br>**250 mg**<br>**(N=599)** | **Placebo**<br>**(N=605)** |
| --- | --- | --- | --- |
| Patients with any adverse event — no. (%) | 267 (44.2) | 321 (53.6) | 150 (24.8) |
| Patients with any treatment-related adverse event — no. (%) | 148 (24.5) | 217 (36.2) | 60 (9.9) |
| Patients with any serious adverse event — no. (%)<sup>b</sup> | 1 (0.2) | 0 | 1 (0.2) |
| Patients with adverse events leading to death — no. (%) | 0 | 0 | 0 |
| Patients with adverse events leading to treatment discontinuation — no. (%)<sup>c</sup> | 4 (0.7) | 6 (1.0) | 2 (0.3) |
| Adverse events occurring in $\geq 2\%$ of patients in either group — no. (%) | | | |
| Headache | 13 (2.2) | 20 (3.3) | 14 (2.3) |
| Diarrhea | 6 (1.0) | 9 (1.5) | 12 (2.0) |
| High-density lipoprotein decreased | 188 (31.1) | 231 (38.6) | 23 (3.8) |
| Blood triglycerides increased | 49 (8.1) | 74 (12.4) | 32 (5.3) |
| Blood bilirubin increased | 36 (6.0) | 56 (9.3) | 6 (1.0) |
| Blood cholesterol decreased | 20 (3.3) | 28 (4.7) | 3 (0.5) |
| Bilirubin conjugated increased | 15 (2.5) | 20 (3.3) | 3 (0.5) |
| Blood creatine phosphokinase increased | 14 (2.3) | 8 (1.3) | 11 (1.8) |
| Blood lactate dehydrogenase increased | 6 (1.0) | 15 (2.5) | 6 (1.0) |
| Aspartate aminotransferase increased | 4 (0.7) | 9 (1.5) | 12 (2.0) |
| Treatment-related adverse events occurring in $\geq 2\%$ of patients in either group — no. (%) | | | |
| Headache | 4 (0.7) | 13 (2.2) | 2 (0.3) |
| High-density lipoprotein decreased | 111 (18.4) | 157 (26.2) | 9 (1.5) |
| Blood triglycerides increased | 16 (2.6) | 37 (6.2) | 17 (2.8) |
| Blood bilirubin increased | 17 (2.8) | 35 (5.8) | 3 (0.5) |
| Blood cholesterol decreased | 8 (1.3) | 12 (2.0) | 1 (0.2) |

<sup>a</sup> Data for all patients who received at least one ensitrelvir or placebo dose are shown.522 <sup>b</sup> Both serious adverse events were judged as not treatment related and recovered with appropriate intervention.
<sup>c</sup> Among the adverse events leading to treatment discontinuation, mild eczema and mild vomiting in one patient each in the ensitrelvir 125-mg group, mild rash and moderate rash in one patient each in the ensitrelvir 250-mg group, and moderate hypoesthesia and mild muscular weakness in one patient in the placebo group were judged as treatment related by the investigator. All treatment-related adverse events leading to treatment discontinuation were deemed to be resolving or resolved after study discontinuation.

Serious adverse events were observed in two patients (heavy menstrual bleeding in one patient in the ensitrelvir 125-mg group on day 14 and acute cholecystitis in one patient in the placebo group on day 9), both of which were judged as not treatment related. A decrease in high-density lipoprotein levels was the most frequently reported adverse event (31.1%, 38.6%, and 3.8% in the ensitrelvir 125-mg, ensitrelvir 250-mg, and placebo groups, respectively) and the most common treatment-related adverse event (18.4%, 26.2%, and 1.5% in the ensitrelvir 125-mg, ensitrelvir 250-mg, and placebo groups, respectively) in both ensitrelvir groups. Adverse events leading to treatment discontinuation were recorded in 12 patients. All treatment-related adverse events leading to treatment discontinuation were deemed to be resolving or resolved after discontinuation of the study. No adverse events leading to death were reported (Table 3).

## Discussion

This phase 3 trial was conducted during the SARS-CoV-2 omicron BA.2 epidemic to assess the efficacy and safety of 5-day, once-daily, oral ensitrelvir for mild-to-moderate COVID-19, using the ensitrelvir 125-mg group as the efficacy analysis population.^20^ More than 90% of the patients had received ≥2 doses of the SARS-CoV-2 vaccine, and approximately 70% of the participants did not have risk factors for severe disease. Treatment with ensitrelvir 125 mg once daily (375 mg on day 1) vs placebo showed a significant difference in the time to resolution of the five typical symptoms of the omicron variant infection in patients randomized in less than 72 hours of disease onset. Moreover, a significant reduction compared with placebo was observed in the two key secondary end points representing antiviral efficacy. In contrast to the results in patients randomized in less than 72 hours of disease onset, no significant difference between ensitrelvir 125 mg and placebo was observed in the time to resolution of the five COVID-19 symptoms in patients randomized within 120 hours, suggesting the importance of early treatment initiation. Ensitrelvir was generally well tolerated, and the most commonly observed adverse event, a decrease in high-density lipoprotein level, is consistent with the findings of previous clinical studies of ensitrelvir.^18,19,23^

Previous clinical trials on oral antivirals for COVID-19 were conducted during the SARS-CoV-2 Delta epidemic.^8,9^ These trials involved unvaccinated, high-risk adults with COVID-19 and demonstrated a reduction in the risk of hospitalization or death as the primary end point.^8,9^ However, given the administration of vaccinations against COVID-19 and the nature of the infection caused by the omicron variant, these end points could no longer be a viable option. Indeed, published studies report a significant reduction in the risk of death, hospitalization, or severe disease with the omicron vs the Delta infection.^3,4,24^ Moreover, real-world studies suggest limited effectiveness of these antivirals to high-risk, vaccinated adults or patients infected with the omicron variant in reducing the risk of severe COVID-19.^25,26^ In the current SCORPIO-SR trial, none of the patients reported mechanical ventilation or death, and only one patient each in the ensitrelvir 250-mg and placebo groups required COVID-19–related hospitalization or equivalent recuperation during the study period (data not shown).

The symptom-based efficacy end points are suggested by the FDA as an alternative goal in the development of antiviral treatment options for COVID-19 caused by recent SARS-CoV-2 variants.^21^ The phase 2/3 EPIC-SR study employed self-reported, sustained alleviation of all targeted COVID-19 symptoms for four days as a novel primary end point to assess the efficacy of nirmatrelvir with ritonavir.^27^ Although a shortened time to sustained alleviation and resolution of COVID-19 signs/symptoms was demonstrated in non-hospitalized adults with COVID-19 at high risk of severe disease (EPIC-HR study),^28^ the EPIC-SR study did not show a meaningful difference in the time to sustained symptom alleviation through day 28.^29^ The current SCORPIO-SR study met the primary end point and demonstrated clinical efficacy of ensitrelvir in the resolution of COVID-19 symptoms (sustained for at least 24 hours) irrespective of risk factors for severe disease. Sustained resolution or alleviation of self-assessed symptoms can be considered a valid and clinically meaningful efficacy end point for COVID-19 treatment.

The major limitation of this study is that it was conducted only in Asian countries, with a limited number of non-Asian patients. The efficacy and safety of ensitrelvir in various patient populations should be further assessed in daily clinical settings.

In conclusion, ensitrelvir treatment vs placebo demonstrated a reduction in the time to resolution of the five typical COVID-19 symptoms in patients treated in less than 72 hours of disease onset. The results also showed favorable antiviral efficacy of ensitrelvir. No new safety concerns were identified, and ensitrelvir was well tolerated.

## Data Availability

Shionogi & Co., Ltd. is committed to disclosing the synopses and results of its clinical trials and sharing the clinical trial data with researchers on reasonable request. For further details, please refer to the websites of Shionogi & Co., Ltd. (https://www.shionogi.com/shionogi/global/en/company/policies/shionogi-group-clinical-trial-data-transparency-policy.html) and Vivli (https://vivli.org/).

## Author Contributions

Dr Uehara had full access to all the data in the study and takes responsibility for the integrity of the data and the accuracy of the data analysis. Concept and design: Hiroshi Yotsuyanagi, Norio Ohmagari, Yohei Doi, Takumi Imamura, Takuhiro Sonoyama, Genki Ichihashi, Takao Sanaki, Yuko Tsuge, Takeki Uehara, and Hiroshi Mukae.

Acquisition, analysis, or interpretation of data: Hiroshi Yotsuyanagi, Norio Ohmagari, Yohei Doi, Masaya Yamato, Nguyen Hoang Bac, Bong Ki Cha, Takumi Imamura, Takuhiro Sonoyama, Genki Ichihashi, Takao Sanaki, Yuko Tsuge, Takeki Uehara, and Hiroshi Mukae. Drafting of the manuscript: Takumi Imamura, Takuhiro Sonoyama, Genki Ichihashi, Takao Sanaki, Yuko Tsuge, Takeki Uehara.

Critical revision of the manuscript for important intellectual content: Hiroshi Yotsuyanagi, Norio Ohmagari, Yohei Doi, Masaya Yamato, Nguyen Hoang Bac, Bong Ki Cha, Takumi Imamura, Takuhiro Sonoyama, Genki Ichihashi, Takao Sanaki, Yuko Tsuge, Takeki Uehara, and Hiroshi Mukae.

Statistical analysis: Takumi Imamura.

Administrative, technical, or material support: Genki Ichihashi, Takao Sanaki, Yuko Tsuge. Supervision: Takeki Uehara, and Hiroshi Mukae.

## Meeting presentations

The work reported herein was presented in part at the joint congress of the 92nd Annual Meeting of the Japanese Association for Infectious Diseases, Western Japan Branch/the 65th Annual Meeting of the Japanese Association for Infectious Diseases, Central Japan Branch/the 70th Annual Meeting of the Japanese Society of Chemotherapy, Western Japan Branch (November 2022).

## Additional Contributions

The authors and research team thank all the patients involved in this study and Masahiro Kinoshita, Manami Yoshida, and Satoshi Kojima (Shionogi & Co., Ltd.) for preparing technical support documents and an earlier version of the manuscript draft. Support for data management was provided by EPS Corporation and funded by Shionogi & Co., Ltd. Medical writing support was provided by Mami Hirano, M.Sc., of Cactus Life Sciences (part of Cactus Communications), and funded by Shionogi & Co., Ltd.

