## Supplement for "Efficacy and Safety of 5-Day Oral Ensitrelvir for Patients With Mild-to-Moderate COVID-19: The SCORPIO-SR Randomized Clinical Trial"


**List of Investigators**

**eMethods**

**eFigure 1.** Time to First Negative SARS-CoV-2 Viral Titer (Modified Intention-to-Treat Population)

**eFigure 2.** SARS-CoV-2 Viral RNA Levels up to Day 21 (Intention-to-Treat Population)

**eFigure 3.** SARS-CoV-2 Viral Titer up to Day 21 (Modified Intention-to-Treat Population)

**eTable 1.** Demographic and Clinical Characteristics of the Patients at Baseline (Safety Analysis Population)

**eTable 2.** Representativeness of Trial Participants

**eTable 3.** Details of Risk Factors for Severe Disease at Baseline (Intention-to-Treat Population)

**eTable 4.** Change from Baseline to Day 4 in SARS-CoV-2 Viral RNA Levels, Time to Resolution of the 12 COVID-19 Symptoms, and Time to Resolution of the 14 COVID-19 Symptoms (Intention-to-Treat Population, Patients Randomized Within 120 Hours of Disease Onset)


This supplemental material has been provided by the authors to give readers additional information about their work.

#### List of Investigators

The following investigators participated in the SCORPIO-SR study and enrolled at least one patient.

| Name | Institute | Location | Institutional review board |
|---|---|---|---|
| **Japan** | | | |
| Fujii, Shigeru | Fukuoka Shinmizumaki Hospital | Onga-gun, Fukuoka, Japan | Adachi Kyosai Hospital Institutional Review Board |
| Fujimaki, Yutaka | Fujimaki Ent Clinic | Ichikawa City, Chiba, Japan | Yoga Allergy Clinic Institutional Review Board |
| Fukuda, Akira | Tokyo Metropolitan Toshima Hospital | Itabashi-ku, Tokyo, Japan | Tokyo Metropolitan Health and Hospitals Toshima Hospital Institutional Review Board |
| Furuichi, Motohiko | Edogawa Medicare Hospital | Edogawa-ku, Tokyo, Japan | Sugiura Clinic Institutional Review Board |
| Haji, Yoichiro | Kojunkai Daido Medicine | Nagoya City, Aichi, Japan | Daido Hospital Institutional Review Board |
| Harada, Toshiyuki | Japan Community Health care Organization Hokkaido Hospital | Sapporo City, Hokkaido, Japan | Review Board of Japan Community Health care Organization Hokkaido Hospital |
| Hayashi, Masamichi | Fujita Health University Okazaki Medical Center | Okazaki City, Aichi, Japan | The Central Institutional Review Board for the Fujita Health University Hospitals |
| Hayashi, Shinichiro | Medical Corporation Kouhoukai Takagi Hospital | Okawa City, Fukuoka, Japan | Review Board of Human Rights and Ethics for Clinical Studies Institutional Review Board |
| Hirai, Yuji | Tokyo Medical University Hachioji Medical Center | Hachioji City, Tokyo, Japan | Tokyo Medical University Hachioji Medical Center Institutional Review Board |
| Hori, Takaki | Kamagaya General Hospital | Kamagaya City, Chiba, Japan | Tokushukai Group Institutional Review Board |
| Igarashi, Tomofumi | Social Medical Corporation Keiwakai Nishioka Hospital | Sapporo City, Hokkaido, Japan | Sapporo Medical Association’s Institutional Review Board |
| Kamezawa, Takashi | Kamezawa Clinic | Kasugai City, Aichi, Japan | Sugiura Clinic Institutional Review Board |
| Kamiya, Uguri | Kaiseikai Kita Shin Yokohama Internal Medicine Clinic | Yokohama City, Kanagawa, Japan | Yoga Allergy Clinic Institutional Review Board |
| Kaneda, Satoru | National Hospital Organization Chiba Medical Center | Chiba City, Chiba, Japan | National Hospital Organization Chiba Medical Center Institutional Review Board |
| Kasamatsu, Yu | University Hospital Kyoto Prefectural University of Medicine | Kyoto City, Kyoto, Japan | University Hospital Kyoto Prefectural University of Medicine Institutional Review Board |
| Kawamura, Naohisa | Japan Organization of Occupational Health and Safety Osaka Rosai Hospital | Sakai City, Osaka, Japan | Osaka Rosai Hospital Institutional Review Board |
| Kisohara, Akira | Kasukabe Medical Center | Kasukabe City, Saitama, Japan | Sugiura Clinic Institutional Review Board |
| Kodaira, Makoto | Kodaira Hospital | Toda City, Saitama, Japan | Sugiura Clinic Institutional Review Board |

| | | | |
|---|---|---|---|
| Konno, Satoshi | Hokkaido University Hospital | Sapporo City, Hokkaido, Japan | Hokkaido University Hospital Institutional Review Board |
| Lee, Woon Joo | Lee's Clinic | Osaka City, Osaka, Japan | Yoga Allergy Clinic Institutional Review Board |
| Matono, Takashi | Aso Iizuka Hospital | Iizuka City, Fukuoka, Japan | Aso Iizuka Hospital Institutional Review Board |
| Murayama, Masanori | Matsunami General Hospital | Hashima-gun, Gifu, Japan | Adachi Kyosai Hospital Institutional Review Board |
| Nakagawa, Hidemitsu | Nozaki Tokushukai Hospital | Daito City, Osaka, Japan | Tokushukai Group Institutional Review Board |
| Nakamura, Fukumi | Tokyo Metropolitan Bokutoh Hospital | Sumida-ku, Tokyo, Japan | Tokyo Metropolitan Bokutoh Hospital Institutional Review Board |
| Nozaki, Minoru | Swing NOZAKI Clinic | Musashino City, Tokyo, Japan | Sugiura Clinic Institutional Review Board |
| Ogata, Tomoyuki | JA Toride Medical Center | Toride City, Ibaraki, Japan | Review Board of Human Rights and Ethics for Clinical Studies Institutional Review Board |
| Ogihara, Shoji | Takinogawa Hospital | Kita-ku, Tokyo, Japan | Yoga Allergy Clinic Institutional Review Board |
| Ohmagari, Norio | Center Hospital of the National Center for Global Health and Medicine | Shinjuku-ku, Tokyo, Japan | Center Hospital of the National Center for Global Health and Medicine Institutional Review Board |
| Ono, Ryuta | Kanagawa Himawari Clinic | Kawasaki City, Kanagawa, Japan | Adachi Kyosai Hospital Institutional Review Board |
| Ota, Kazue | Minna no Tennocho Clinic | Yokohama City, Kanagawa, Japan | Adachi Kyosai Hospital Institutional Review Board |
| Ota, Kazue | Yaguchi Midori Clinic | Ota-ku, Tokyo, Japan | Adachi Kyosai Hospital Institutional Review Board |
| Owan, Isoko | National Hospital Organization Okinawa National Hospital | Ginowan City, Okinawa, Japan | Okinawa National Hospital Clinical Trial Review Committee |
| Sagara, Hironori | SHOWA University East Hospital | Shinagawa-ku, Tokyo, Japan | Showa University Hospital Institutional Review Board |
| Shimizu, Hidefumi | Japan Community Health care Organization Tokyo Shinjuku Medical Center | Shinjuku-ku, Tokyo, Japan | Sugiura Clinic Institutional Review Board |
| Shimizu, Masatoshi | National Hospital Organization Kobe Medical Center | Kobe City, Hyogo, Japan | National Hospital Organization Kobe Medical Center Institutional Review Board |
| Suzuki, Hiromichi | University of Tsukuba Hospital | Tsukuba City, Ibaraki, Japan | Institutional Review Board, University of Tsukuba Hospital |
| Tachikawa, Natsuo | Yokohama Municipal Citizen's Hospital | Yokohama City, Kanagawa, Japan | Yokohama Municipal Citizen's Hospital Institutional Review Board |
| Tajima, Yasuhisa | Hamamatsu Medical Center | Hamamatsu City, Shizuoka, Japan | Hamamatsu Medical Center Institutional Review Board |
| Takahashi, Satoshi | Sapporo Medical University Hospital | Sapporo City, Hokkaido, Japan | Sapporo Medical University Hospital Institutional Review Board |
| Tashiro, Naotaka | Tashiro Thyroid Clinic | Fukuoka City, Fukuoka, Japan | Adachi Kyosai Hospital Institutional Review Board |

| | | | |
|---|---|---|---|
| Tsushima, Kenji | IUHW Narita Hospital | Narita City, Chiba, Japan | International University of Health and Welfare and Institutional Review Board |
| Umezawa, Yoshihiro | Den-en-chofu Family Clinic | Ota-ku, Tokyo, Japan | Kobori Central Clinical Research Ethics Committee |
| Yada, Shinichiro | Onga Nakama Medical Association Onga Hospital | Onga-gun, Fukuoka, Japan | Sugiura Clinic Institutional Review Board |
| Yamada, Kota | Tsuchiura Beryl Clinic | Tsuchiura City, Ibaraki, Japan | Sugiura Clinic Institutional Review Board |
| Yamato, Masaya | Rinku General Medical Center | Izumisano City, Osaka, Japan | Rinku General Medical Center Institutional Review Board |
| Yamato, Tsuyoshi | Kouwakai Kouwa Clinic | Toshima-ku, Tokyo, Japan | Sugiura Clinic Institutional Review Board |
| **Vietnam** | | | |
| Do, Van Dung | University of Medicine and Pharmacy at Ho Chi Minh city | District 5, Ho Chi Minh city, Vietnam | Ethics Committee in Biomedical Research of University of Medicine and Pharmacy in Ho Chi Minh city |
| Pham, Thi Van Anh | Hanoi Medical University | Dong Da District, Hanoi, Vietnam | Hanoi Medical University Institutional Ethical Review Board |
| **South Korea** | | | |
| Cha, Bongki | Chung-Ang University Health Care System Hyundae Hospital | Namyangju-si, Gyeonggi-do, South Korea | Chung-Ang University Healthcare System Hyundae Hospital Institutional Review Board |
| Han, Sang Hoon | Gangnam Severance Hospital | Gangnam-gu, Seoul, South Korea | Gangnam Severance Hospital Institutional Review Board |
| Heo, Eun Young | SMG-SNU Boramae Medical Center | Dongjak-gu, Seoul, South Korea | Seoul Metropolitan Government Seoul National University Boramae Medical Center Institutional Review Board |
| Kang, Yumin | Myongji Hospital | Goyang-si, Gyeonggi-do, South Korea | Myongji Hospital Institutional Review Board |
| Kwak, Yee Gyung | Inje University Ilsan Paik Hospital | Goyang-si, Gyeonggi-do, South Korea | Inje University Ilsan Paik Hospital Institutional Review Board |
| Lee, Eung Gu | The Catholic University of Korea Bucheon St. Mary's Hospital | Bucheon-si, Gyeonggi-do, South Korea | The Catholic Medical Center Central Institutional Review Board |
| Lee, Heayon | The Catholic University of Korea Eunpyeong St. Mary's Hospital | Eunpyeong-gu, Seoul, South Korea | The Catholic Medical Center Central Institutional Review Board |
| Lee, Jin Soo | Inha University Hospital | Jung-gu, Incheon, South Korea | Inha University Hospital Institutional Review Board |
| Lee, Mi Suk | Kyung Hee University Medical Center | Dongdaemun-gu, Seoul, South Korea | Kyung Hee University Hospital, Institutional Review Board |
| Park, Jinsik | Incheon Sejong Hospital | Gyeyang-gu, Incheon, South Korea | Incheon Sejong Hospital Institutional Review Board |
| Park, Seong Yeon | Dongguk University Ilsan Hospital | Goyang-si, Gyeonggi-do, South Korea | Dogguk University Ilsan Hospital Institutional Review Board |

| | | | |
|---|---|---|---|
| Park, Yoon Soo | Yongin Severance Hospital | Yongin-si, Gyeonggi-do, South Korea | YONGIN SEVERANCE HOSPITAL Institutional Review Board |
| Shi, Hye Jin | Gachon University Gil Medical Center | Namdong-gu, Incheon, South Korea | Gachon University Gil Medical Center Institutional Review Board |

#### eMethods

##### Inclusion and Exclusion Criteria

Patients aged 12 to <70 years who tested positive for severe acute respiratory syndrome coronavirus 2 (SARS-CoV-2) within 120 hours prior to randomization and had a time from the onset of coronavirus disease 2019 (COVID-19) symptom (at least one of the 14 symptoms listed in "COVID-19 Symptom Scores" below) to randomization of ≤120 hours were eligible for study enrollment. SARS-CoV-2 tests were performed using a nucleic acid detection test using a nasopharyngeal swab, nasal swab, or saliva (qualitative/quantitative reverse transcription-polymerase chain reaction [RT-PCR] test or an isothermal nucleic acid amplification method [e.g., the loop-mediated isothermal amplification method or the transcription-mediated amplification method]); a quantitative antigen test using a nasopharyngeal swab, nasal swab, or saliva; or a qualitative antigen test using a nasopharyngeal or nasal swab. To avoid excessive drug exposure, patients aged <18 years should have recorded a body weight of ≥40 kg at enrollment. Patients with mild-to-moderate COVID-19 were defined as those who have had at least one moderate or severe symptom among the 12 COVID-19 symptoms at enrollment (excluding symptoms present prior to COVID-19 onset) or those who have at least one moderate or severe existing symptom or symptoms presenting prior to COVID-19 onset that was considered to have worsened at baseline (refer to "COVID-19 Symptom Scores" below for the details of symptom severity assessments).
The key exclusion criteria included the following: an awake oxygen saturation of ≤93% (room air); supplemental oxygen requirement; anticipated COVID-19 exacerbation within 48 hours of randomization in the opinion of the investigator; suspected active and systemic infections other than COVID-19 requiring treatment; current or chronic history of moderate or severe liver disease, known hepatic or biliary abnormalities (except for Gilbert's syndrome or asymptomatic gallstones), or moderate-to-severe kidney disease; pregnancy, possible pregnancy, or lactation; and blood donation (≥400 mL within 12 weeks or ≥200 mL within 4 weeks prior to enrollment). Patients who had used drugs for SARS-CoV-2 infection within 7 days prior to randomization, a strong cytochrome P450, family 3, subfamily A (CYP3A) inhibitor or inducer within 14 days prior to randomization, or St. John's wort products within 14 days prior to randomization were also excluded.

##### COVID-19 Symptom Scores

| Symptom | Symptom rating | Five symptoms | 12 symptoms | 14 symptoms |
|---|---|---|---|---|
| 1. Stuffy or runny nose | None=0<br>Mild=1<br>Moderate=2<br>Severe=3 | ● | ● | ● |
| 2. Sore throat | | ● | ● | ● |
| 3. Shortness of breath (difficulty breathing) | | | ● | ● |
| 4. Cough | | ● | ● | ● |
| 5. Low energy or tiredness | | ● | ● | ● |
| 6. Muscle or body aches | | | ● | ● |
| 7. Headache | | | ● | ● |
| 8. Chills or shivering | | | ● | ● |
| 9. Feeling hot or feverish | | ● | ● | ● |
| 10. Nausea (feeling like you want to throw up) | | | ● | ● |
| 11. Vomiting (throwing up) | | | ● | ● |
| 12. Diarrhea (loose or watery stools) | | | ● | ● |
| 13. Sense of smell | Same as usual=0<br>Less than usual=1<br>No sense of smell/taste=2 | | | ● |
| 14. Sense of taste | | | | ● |

##### Treatment

Patients received an oral once-daily dosage of ensitrelvir 125 mg, ensitrelvir 250 mg, or matching placebo tablets. Two types of matching placebo tablets were used: placebo-B, which was identical in appearance and packaging to ensitrelvir 125 mg, and placebo-D, which was identical in appearance and packaging to ensitrelvir 250 mg. Patients randomized to the ensitrelvir 125-mg group received three tablets of ensitrelvir 125 mg (i.e., 375 mg in total as a loading dose) and three placebo-D tablets on day 1. On days 2 to 5, one tablet each of ensitrelvir 125 mg and placebo-D was administered. Similarly, patients randomized to the ensitrelvir 250-mg group received three tablets of ensitrelvir 250 mg (i.e., 750 mg in total as a loading dose) and three placebo-B tablets on day 1, followed by one tablet each of ensitrelvir 250 mg and placebo-B administered on days 2 to 5. Patients assigned to the placebo group received three tablets each of placebo-D and placebo-B on day 1, followed by one tablet each of placebo-D and placebo-B administered on days 2 to 5. In view of the CYP3A inhibitory effect of ensitrelvir, intake of any foods or beverages containing grapefruit or Seville oranges or use of St. John's wort products was prohibited until day 5.

**Study Assessments**

***COVID-19 Symptom Assessments***

Patients self-assessed the severity of 14 COVID-19 symptoms (refer to "COVID-19 Symptom Scores" above) twice daily, in the morning and evening, on days 1 to 9 and once daily in the evening from days 10 to 21. Symptom assessments were postponed until 4 hours after drug administration in patients taking acetaminophen for antipyretic or analgesic purposes. The time to resolution of the five COVID-19 symptoms was defined as the time from the start of the study intervention to the resolution of all five symptoms. Patients were considered to have achieved the primary end point if all five symptoms remained resolved for ≥24 hours. Resolution of symptoms was defined as follows: (1) for pre-existing symptoms that were present before the onset of COVID-19 and considered by the patient to have worsened at baseline, severe symptoms at baseline must have improved to moderate or better, moderate symptoms at baseline must have improved to mild or better, and mild symptoms at baseline must have remained mild or better (no symptoms) and (2) for pre-existing symptoms that were present before the onset of COVID-19 and considered by the patient not to have worsened at baseline, severe symptoms at baseline must have remained severe or improved, moderate symptoms at baseline must have remained moderate or improved, and mild symptoms at baseline must have remained mild or better (no symptoms). Symptoms other than those mentioned above (those not occurring before the onset of COVID-19 or those that occur at or after baseline) must have been completely resolved. The time to resolution of the 12 and 14 COVID-19 symptoms was defined in a similar manner, i.e., the time from the start of the study intervention to the resolution of all the symptoms.

***Virologic Assessments***

Nasopharyngeal swabs collected from patients by the investigator or his/her designee were used to measure the SARS-CoV-2 viral RNA levels and viral titers. SARS-CoV-2 viral titers and viral RNA levels were centrally measured at ViroClinics (Rotterdam, Netherlands). RT-PCR tests were performed to determine the presence or absence of SARS-CoV-2 viral RNA.

***Safety Assessments***

In addition to the reporting of adverse events, laboratory tests, vital sign measurements, and electrocardiography were performed throughout the study period. Pregnancy tests were performed on women with childbearing potential on day 1 (before drug administration) and day 28 (or study discontinuation). Additional pregnancy tests were permitted at the discretion of the investigator. All safety data were evaluated by an independent Data and Safety Monitoring Board.

**Statistical Analysis**

The primary end point (time to resolution of the five COVID-19 symptoms), key secondary end point 1 (change from baseline in SARS-CoV-2 viral RNA level on day 4), and other secondary end points (time to resolution of the 12 COVID-19 symptoms, time to resolution of the 14 COVID-19 symptoms, and SARS-CoV-2 viral RNA up to day 21) were assessed in the intention-to-treat population. The intention-to-treat population comprised all patients who tested positive for SARS-CoV-2 viral RNA at baseline, as confirmed by an RT-PCR test based on the nasopharyngeal swab sample. The key secondary end point 2 (time to first negative SARS-CoV-2 viral titer) and other secondary end points (SARS-CoV-2 viral titer up to day 21) were assessed in the modified intention-to-treat population. The modified intention-to-treat population comprised all patients who tested positive for SARS-CoV-2 viral RNA and who had detectable SARS-CoV-2 viral titer at baseline. All safety assessments were performed in the safety analysis population comprising all patients who received at least one ensitrelvir or placebo dose.

For the analyses in the primary analysis population (patients randomized in less than 72 hours of disease onset in the ensitrelvir 125-mg group), the time to resolution of the five COVID-19 symptoms was compared with the placebo group using a Peto-Prentice generalized Wilcoxon test stratified by SARS-CoV-2 vaccination history (yes or no). The change from baseline in the SARS-CoV-2 viral RNA level on day 4 was compared with that in the placebo group using an analysis of covariance (ANCOVA) model; baseline SARS-CoV-2 viral RNA and SARS-CoV-2 vaccination history (yes or no) were included as covariates. The time to the first negative SARS-

CoV-2 viral titer was compared with that of the placebo group using a log-rank test stratified by SARS-CoV-2 vaccination history (yes or no). All statistical comparisons were performed at a two-sided significance level of .05. For the analyses of patients randomized within 120 hours of disease onset, the time from onset to randomization (<72 hours or ≥72 hours) was added as a stratification factor or covariate.

#### eFigure 1. Time to First Negative SARS-CoV-2 Viral Titer (Modified Intention-to-Treat Population)

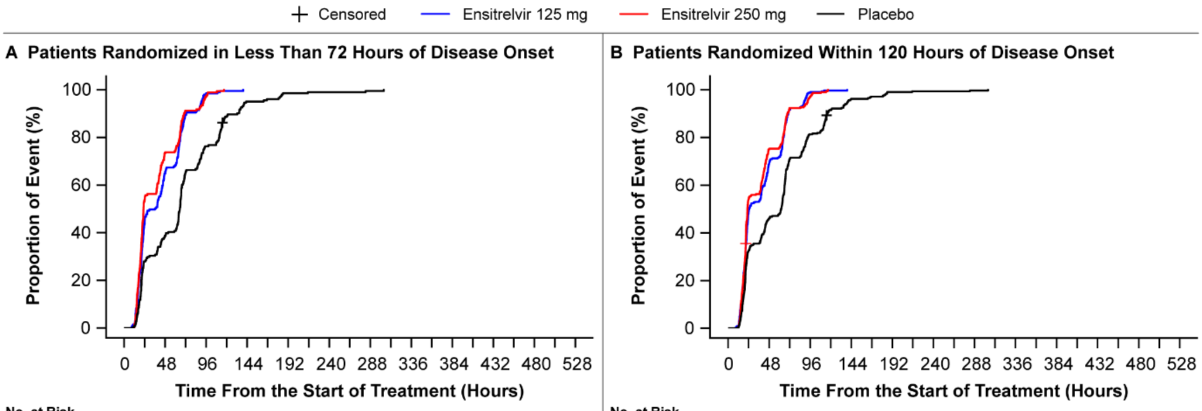


**A** No. at Risk

| | 0 | 48 | 96 | 144 | 192 | 240 | 288 | 336 | 384 | 432 | 480 | 528 |
|---|---|---|---|---|---|---|---|---|---|---|---|---|
| Ensitrelvir 125 mg | 199 | 70 | 3 | 0 | 0 | 0 | 0 | 0 | 0 | 0 | 0 | 0 |
| Ensitrelvir 250 mg | 183 | 48 | 5 | 0 | 0 | 0 | 0 | 0 | 0 | 0 | 0 | 0 |
| Placebo | 211 | 129 | 50 | 10 | 3 | 2 | 1 | 0 | 0 | 0 | 0 | 0 |

**B** No. at Risk

| | 0 | 48 | 96 | 144 | 192 | 240 | 288 | 336 | 384 | 432 | 480 | 528 |
|---|---|---|---|---|---|---|---|---|---|---|---|---|
| Ensitrelvir 125 mg | 313 | 97 | 3 | 0 | 0 | 0 | 0 | 0 | 0 | 0 | 0 | 0 |
| Ensitrelvir 250 mg | 301 | 74 | 9 | 0 | 0 | 0 | 0 | 0 | 0 | 0 | 0 | 0 |
| Placebo | 327 | 176 | 61 | 12 | 3 | 2 | 1 | 0 | 0 | 0 | 0 | 0 |

The analysis was performed for all patients who tested positive for SARS-CoV-2 viral RNA, with a detectable SARS-CoV-2 viral titer at baseline. Patients randomized in less than 72 hours of disease onset in the ensitrelvir 125-mg group are defined as the primary analysis population. A stratified log-rank test was applied to test the statistical significance vs placebo. The test was stratified by SARS-CoV-2 vaccination history (yes or no) for patients randomized in less than 72 hours (panel A) and time from onset to randomization (<72 hours or ≥72 hours) and SARS-CoV-2 vaccination history (yes or no) for patients randomized within 120 hours (panel B).
SARS-CoV-2, severe acute respiratory syndrome coronavirus 2.

#### eFigure 2. SARS-CoV-2 Viral RNA Levels up to Day 21 (Intention-to-Treat Population)

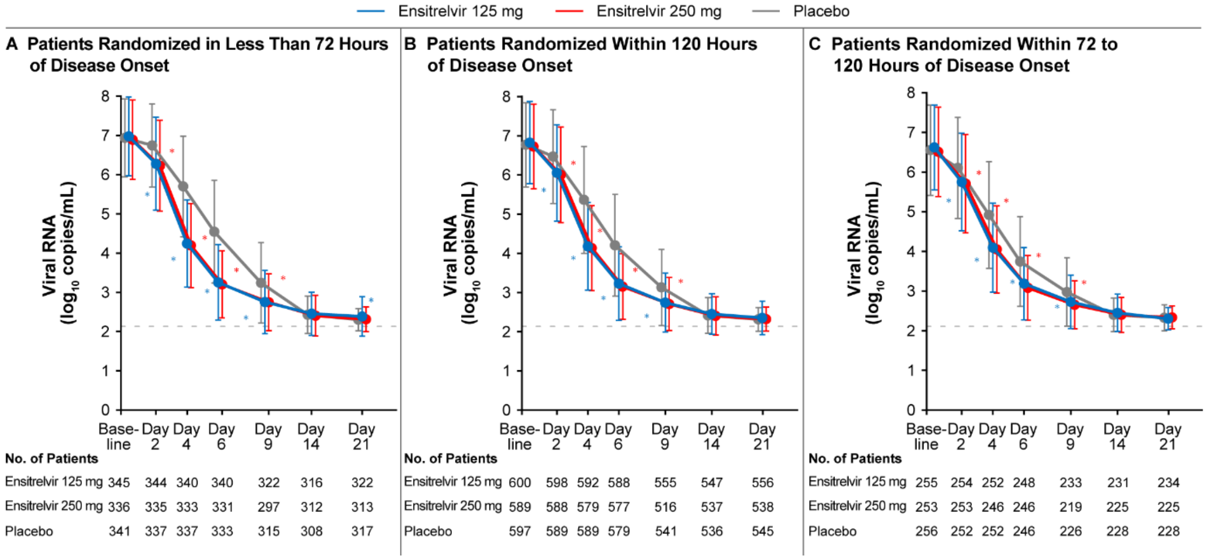


The analysis was performed for all patients who tested positive for SARS-CoV-2 viral RNA at baseline. Patients randomized in less than 72 hours of disease onset in the ensitrelvir 125-mg group were defined as the primary analysis population. The gray dotted line represents the LLoQ for SARS-CoV-2 viral RNA (2.08 $log_{10}$ copies/mL). A viral RNA level lower than the LLoD (negative viral RNA) and that lower than the LLoQ were imputed as 2.27 (lower limit of 95% detection) and 2.08 $log_{10}$ copies/mL, respectively. The covariates used for ANCOVA are baseline SARS-CoV-2 viral RNA and SARS-CoV-2 vaccination history (yes or no) for patients randomized in less than 72 hours (panel A); baseline SARS-CoV-2 viral RNA, time from onset to randomization (<72 hours or ≥72 hours), and SARS-CoV-2 vaccination history (yes or no) for patients randomized within 120 hours (panel B); and baseline SARS-CoV-2 viral RNA and SARS-CoV-2 vaccination history (yes or no) for patients randomized within 72 to 120 hours (panel C).
* P value (ANCOVA) < .05.
ANCOVA, analysis of covariance; LLoD, lower limit of detection; LLoQ, lower limit of quantification; SARS-CoV-2, severe acute respiratory syndrome coronavirus 2.

### eFigure 3. SARS-CoV-2 Viral Titer up to Day 21 (Modified Intention-to-Treat Population)

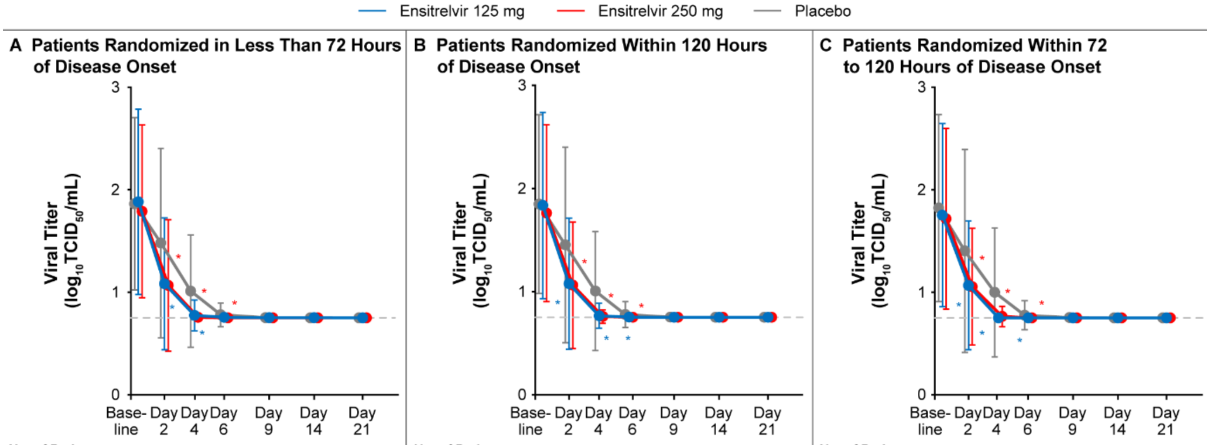


**A** No. of Patients

| | Baseline | Day 2 | Day 4 | Day 6 | Day 9 | Day 14 | Day 21 |
|---|---|---|---|---|---|---|---|
| Ensitrelvir 125 mg | 201 | 196 | 197 | 195 | 194 | 180 | 179 |
| Ensitrelvir 250 mg | 184 | 182 | 180 | 181 | 172 | 163 | 164 |
| Placebo | 212 | 208 | 207 | 208 | 205 | 182 | 189 |

**B** No. of Patients

| | Baseline | Day 2 | Day 4 | Day 6 | Day 9 | Day 14 | Day 21 |
|---|---|---|---|---|---|---|---|
| Ensitrelvir 125 mg | 315 | 310 | 310 | 302 | 300 | 280 | 283 |
| Ensitrelvir 250 mg | 302 | 299 | 294 | 294 | 274 | 264 | 265 |
| Placebo | 331 | 326 | 324 | 320 | 310 | 284 | 294 |

**C** No. of Patients

| | Baseline | Day 2 | Day 4 | Day 6 | Day 9 | Day 14 | Day 21 |
|---|---|---|---|---|---|---|---|
| Ensitrelvir 125 mg | 114 | 114 | 113 | 107 | 106 | 100 | 104 |
| Ensitrelvir 250 mg | 118 | 117 | 114 | 113 | 102 | 101 | 101 |
| Placebo | 119 | 118 | 117 | 112 | 105 | 102 | 105 |

The analysis was performed for all patients who tested positive for SARS-CoV-2 viral RNA, with a detectable SARS-CoV-2 viral titer at baseline. Patients randomized in less than 72 hours of disease onset in the ensitrelvir 125-mg group were defined as the primary analysis population. The gray dotted line represents the LLoD for SARS-CoV-2 viral titer (0.75 $\log_{10}$ $TCID_{50}$/mL). A viral titer lower than the LLoD (negative viral titer) and that lower than the LLoQ were imputed as 0.75 (LLoD) and 1.0 (LLoQ) $\log_{10}$ $TCID_{50}$/mL, respectively. The covariates used for ANCOVA are baseline SARS-CoV-2 viral titer and SARS-CoV-2 vaccination history (yes or no) for patients randomized in less than 72 hours (panel A); baseline SARS-CoV-2 viral titer, time from onset to randomization (<72 hours or ≥72 hours), and SARS-CoV-2 vaccination history (yes or no) for patients randomized within 120 hours (panel B); and SARS-CoV-2 vaccination history (yes or no) for patients randomized within 72 to 120 hours (panel C). * P value (ANCOVA) < .05.
ANCOVA, analysis of covariance; LLoD, lower limit of detection; LLoQ, lower limit of quantification; SARS-CoV-2, severe acute respiratory syndrome coronavirus 2; $TCID_{50}$, 50% tissue-culture infectious dose.

#### eTable 1. Demographic and Clinical Characteristics of the Patients at Baseline (Safety Analysis Population)

| Characteristic | Ensitrelvir 125 mg (N=604) | Ensitrelvir 250 mg (N=599) | Placebo (N=605) |
|---|---|---|---|
| Male sex — no. (%) | 319 (52.8) | 325 (54.3) | 311 (51.4) |
| Mean (SD) age — years | 36.0 (12.6) | 35.9 (12.7) | 35.3 (12.6) |
| Asian race — no. (%) | 602 (99.7) | 597 (99.7) | 603 (99.7) |
| COVID-19 vaccination history — no. (%) | | | |
| ≥1 vaccination | 562 (93.0) | 555 (92.7) | 558 (92.2) |
| ≥2 vaccinations | 554 (91.7) | 551 (92.0) | 549 (90.7) |
| ≥3 vaccinations | 286 (47.4) | 295 (49.2) | 288 (47.6) |
| Prior acetaminophen use — no. (%) | 216 (35.8) | 194 (32.4) | 209 (34.5) |

Data for all patients who received at least one ensitrelvir or placebo dose are shown.
COVID-19, coronavirus disease 2019; SD, standard deviation.

#### eTable 2. Representativeness of Trial Participants

| **Disease under investigation** | **COVID-19** |
|---|---|
| Special considerations related to | |
| Sex and gender | COVID-19 affects men and women with similar frequency. Data on gender identity and COVID-19 remain limited. |
| Age | In adults, the prevalence of COVID-19 is generally similar across age groups. The elderly population is at an increased risk of mortality. |
| Race or ethnic group | COVID-19 affects people of all races and ethnicities. |
| Geography | The COVID-19 pandemic poses a global threat, and its clinical burden is not limited to a specific country or region. |
| Other considerations | The characteristics of SARS-CoV-2 are highly variable among variants. Omicron, the circulating variant of concern as of January 2023, is characterized by disease of mild severity. However, immune-evasive omicron subvariants are being newly identified globally. Despite the worldwide initiatives for vaccination against SARS-CoV-2 infection, the vaccination status greatly varies among regions; as of January 2023, South-East Asia is ranked third among the six global regions defined by the World Health Organization in terms of the number of persons fully vaccinated per 100 population.[1] |
| Overall representativeness of this trial | The participants in the present trial demonstrated the expected ratio of men to women. Biologic sex was self-reported by trial participants, but gender identity was not collected as it was not a part of the regulatory or legal requirements for conducting clinical trials in participating countries. Approximately half of the patients were men across treatment groups. Patient age was approximately 35 years on average in all treatment groups. The present trial was designed to enroll patients from Japan, Vietnam, and South Korea; the trial participants were enrolled from these three countries (Japan [53%], Vietnam [38%], and South Korea [9%]). More than 87% of the participants were classified under “not Hispanic or Latino ethnicity,” and almost all patients were Asian. The present trial was conducted during the SARS-CoV-2 omicron epidemic and designed to enroll patients with mild-to-moderate COVID-19 irrespective of risk factors for severe disease. A majority (>90%) of patients had received ≥2 doses of the SARS-CoV-2 vaccine, and >85% of the patients were infected with the omicron BA.1 or BA.2 subvariant. Approximately 70% of the participants were those without risk factors for severe disease. |

COVID-19, coronavirus disease 2019; SARS-CoV-2, severe acute respiratory syndrome coronavirus 2.

1. World Health Organization. WHO Coronavirus (COVID-19) Dashboard: Situation by Region, Country, Territory & Area. https://covid19.who.int/table. Accessed June 15, 2023.

**eTable 3. Details of Risk Factors for Severe Disease at Baseline (Intention-to-Treat Population)**

| Characteristic | Patients Randomized in Less Than 72 Hours of Disease Onset | | | Patients Randomized Within 120 Hours of Disease Onset | | |
|---|---|---|---|---|---|---|
| | Ensitrelvir 125 mg (N=347)[a] | Ensitrelvir 250 mg (N=340) | Placebo (N=343) | Ensitrelvir 125 mg (N=603) | Ensitrelvir 250 mg (N=595) | Placebo (N=600) |
| Patients with any risk factors — no. (%) | 107 (30.8) | 90 (26.5) | 89 (25.9) | 174 (28.9) | 167 (28.1) | 152 (25.3) |
| Age ≥65 years | 2 (0.6) | 2 (0.6) | 2 (0.6) | 4 (0.7) | 5 (0.8) | 7 (1.2) |
| BMI ≥30 kg/m$^2$ | 23 (6.6) | 15 (4.4) | 12 (3.5) | 34 (5.6) | 32 (5.4) | 24 (4.0) |
| Cancer | 0 | 3 (0.9) | 0 | 1 (0.2) | 3 (0.5) | 0 |
| Cerebrovascular disease | 0 | 0 | 0 | 0 | 0 | 0 |
| Chronic kidney disease | 0 | 0 | 0 | 0 | 0 | 0 |
| Chronic lung disease | 1 (0.3) | 0 | 1 (0.3) | 2 (0.3) | 0 | 1 (0.2) |
| Chronic liver disease | 0 | 0 | 0 | 0 | 0 | 0 |
| Cystic fibrosis | 0 | 0 | 0 | 0 | 0 | 0 |
| Diabetes mellitus | 4 (1.2) | 6 (1.8) | 5 (1.5) | 7 (1.2) | 8 (1.3) | 7 (1.2) |
| Disabilities | 1 (0.3) | 0 | 0 | 2 (0.3) | 0 | 0 |
| Heart conditions | 0 | 2 (0.6) | 1 (0.3) | 1 (0.2) | 3 (0.5) | 2 (0.3) |
| Hypertension | 22 (6.3) | 10 (2.9) | 14 (4.1) | 34 (5.6) | 28 (4.7) | 20 (3.3) |
| Dyslipidemia | 29 (8.4) | 17 (5.0) | 26 (7.6) | 36 (6.0) | 32 (5.4) | 35 (5.8) |
| Human immunodeficiency virus | 0 | 0 | 0 | 0 | 0 | 0 |
| Mental health disorders | 2 (0.6) | 1 (0.3) | 0 | 4 (0.7) | 4 (0.7) | 3 (0.5) |
| Neurologic conditions | 0 | 0 | 0 | 0 | 0 | 0 |
| Primary immunodeficiencies | 0 | 0 | 0 | 0 | 0 | 0 |
| Smoking | 55 (15.9) | 56 (16.5) | 48 (14.0) | 94 (15.6) | 98 (16.5) | 87 (14.5) |
| Solid organ or hematopoietic cell transplantation | 0 | 0 | 0 | 0 | 0 | 0 |
| Tuberculosis | 0 | 0 | 0 | 0 | 0 | 0 |

| Characteristic | Patients Randomized in Less Than 72 Hours of Disease Onset | | | Patients Randomized Within 120 Hours of Disease Onset | | |
|---|---|---|---|---|---|---|
| | Ensitrelvir 125 mg (N=347)[a] | Ensitrelvir 250 mg (N=340) | Placebo (N=343) | Ensitrelvir 125 mg (N=603) | Ensitrelvir 250 mg (N=595) | Placebo (N=600) |
| Use of corticosteroids or other immunosuppressive medications | 4 (1.2) | 3 (0.9) | 3 (0.9) | 5 (0.8) | 3 (0.5) | 4 (0.7) |

Data for all patients who tested positive for SARS-CoV-2 viral RNA at baseline are shown.
[a] Primary analysis population.
BMI, body mass index; SARS-CoV-2, severe acute respiratory syndrome coronavirus 2.

**eTable 4. Change from Baseline to Day 4 in SARS-CoV-2 Viral RNA Levels, Time to Resolution of the 12 COVID-19 Symptoms, and Time to Resolution of the 14 COVID-19 Symptoms (Intention-to-Treat Population, Patients Randomized Within 120 Hours of Disease Onset)**

| Variable | Ensitrelvir 125 mg (N=603) | Ensitrelvir 250 mg (N=595) | Placebo (N=600) |
|---|---|---|---|
| Change from baseline to day 4 in SARS-CoV-2 viral RNA levels[a] | | | |
| N | 592 | 579 | 589 |
| LS mean (SE) — $\log_{10}$ copies/mL | −2.46 (0.06) | −2.46 (0.06) | −1.26 (0.06) |
| Difference from placebo, LS mean (SE) | −1.20 (0.06) | −1.20 (0.06) | |
| 95% CI | −1.32 to −1.08 | −1.32 to −1.08 | |
| P value | <.001 | <.001 | |
| Time to resolution of the 12 COVID-19 symptoms[b] | | | |
| N | 582 | 577 | 572 |
| Median (95% CI) — hours | 200.0 (179.2 to 241.3) | 192.1 (180.2 to 231.1) | 221.5 (192.2 to 246.5) |
| Difference from placebo, median (95% CI) | −21.5 (−55.1 to 29.3) | −29.4 (−59.1 to 19.8) | |
| P value[c] | .76 | .35 | |
| Time to resolution of the 14 COVID-19 symptoms[d] | | | |
| N | 582 | 577 | 572 |
| Median (95% CI) — hours | 215.9 (187.9 to 246.0) | 212.7 (183.9 to 246.9) | 240.2 (206.3 to 266.5) |
| Difference from placebo, median (95% CI) | −24.2 (−66.5 to 26.3) | −27.4 (−69.4 to 27.1) | |
| P value[c] | .46 | .30 | |

The analysis was performed for all patients who tested positive for SARS-CoV-2 viral RNA at baseline.
[a] Analysis by ANCOVA using baseline SARS-CoV-2 viral RNA, time from onset to randomization (<72 hours or ≥72 hours), and SARS-CoV-2 vaccination history (yes or no) as covariates.
[b] Low energy or tiredness, muscle or body aches, headache, chills or shivering, feeling hot or feverish, stuffy or runny nose, sore throat, cough, shortness of breath (difficulty breathing), nausea, vomiting, and diarrhea.
[c] Based on a Peto-Prentice generalized Wilcoxon test stratified by time from onset to randomization (<72 hours or ≥72 hours) and SARS-CoV-2 vaccination history (yes or no).
[d] Low energy or tiredness, muscle or body aches, headache, chills or shivering, feeling hot or feverish, stuffy or runny nose, sore throat, cough, shortness of breath (difficulty breathing), nausea, vomiting, diarrhea, anosmia, and dysgeusia.
ANCOVA, analysis of covariance; CI, confidence interval; COVID-19, coronavirus disease 2019; LS, least squares; SARS-CoV-2, severe acute respiratory syndrome coronavirus 2; SE, standard error.
